# Evaluating GPT-4-based ChatGPT’s Clinical Potential on the NEJM Quiz

**DOI:** 10.1101/2023.05.04.23289493

**Authors:** Daiju Ueda, Shannon L Walston, Toshimasa Matsumoto, Ryo Deguchi, Hiroyuki Tatekawa, Yukio Miki

**Author notes:** <u>**Corresponding author**</u> Daiju Ueda, MD, **<u>E-mail</u>**. **<u>Funding information</u>** This study is funded by Iida Home Max Co., Ltd. **<u>Data sharing statement</u>** All data generated or analyzed during the study are included in the published paper.

## Abstract

**Background:** GPT-4-based ChatGPT demonstrates significant potential in various industries; however, its potential clinical applications remain largely unexplored.

**Methods:** We employed the New England Journal of Medicine (NEJM) quiz “Image Challenge” from October 2021 to March 2023 to assess ChatGPT’s clinical capabilities. The quiz, designed for healthcare professionals, tests the ability to analyze clinical scenarios and make appropriate decisions. We evaluated ChatGPT’s performance on the NEJM quiz, analyzing its accuracy rate by questioning type and specialty after excluding quizzes which were impossible to answer without images. The NEJM quiz has five multiple-choice options, but ChatGPT was first asked to answer without choices, and then given the choices to answer afterwards, in order to evaluate the accuracy in both scenarios.

**Results:** ChatGPT achieved an 87% accuracy without choices and a 97% accuracy with choices, after excluding 16 image-based quizzes. Upon analyzing performance by quiz type, ChatGPT excelled in the Diagnosis category, attaining 89% accuracy without choices and 98% with choices. Although other categories featured fewer cases, ChatGPT’s performance remained consistent. It demonstrated strong performance across the majority of medical specialties; however, Genetics had the lowest accuracy at 67%.

**Conclusion:** ChatGPT demonstrates potential for clinical application, suggesting its usefulness in supporting healthcare professionals and enhancing AI-driven healthcare.

## Introduction

In recent years, the field of artificial intelligence (AI) has witnessed rapid advancements, particularly in the domain of natural language processing (NLP).^1^ The development of advanced NLP models has revolutionized the way humans interact with computers, enabling machines to better understand and respond to complex linguistic inputs. As AI systems become increasingly intuitive and capable, they present the potential to transform a multitude of industries and improve the quality of life for millions of people worldwide.^1^

The advent of ChatGPT, and specifically the GPT-4 architecture, has resulted in a multitude of applications and research opportunities.^2,3^ GPT-4 has displayed an unparalleled level of language understanding and generation capabilities, far surpassing its predecessors in terms of performance and versatility.^4,5^ Its ability to grasp context, generate coherent and contextually relevant responses, and adapt to a wide range of tasks has made it an invaluable asset for numerous domains. As researchers and industries continue to explore the potential of GPT-4, its role in shaping the future of human-computer interaction becomes increasingly apparent.

Despite the growing prominence of ChatGPT, the exploration of its potential clinical applications remains largely uncharted. There is a significant gap in our understanding of how ChatGPT can be harnessed to support healthcare professionals in their daily practice, aid in clinical decision-making processes, or contribute to patient education and engagement. This lack of knowledge underscores the need for more in-depth investigations into the clinical capabilities of ChatGPT, and the need for exploring its potential to revolutionize healthcare and improve patient outcomes.

To assess the clinical applicability of ChatGPT, we employed the New England Journal of Medicine (NEJM) quiz as a benchmark. This rigorous quiz, designed for healthcare professionals, tests the ability to analyze clinical scenarios, synthesize information, and make appropriate decisions. By analyzing ChatGPT’s performance on the NEJM quiz, we sought to determine its potential to assist clinicians in their daily practice, contribute to the ever-growing field of AI-driven healthcare, and help transform the way healthcare professionals approach decision-making and patient care. This evaluation aims to provide a foundation for future research and development, paving the way for more widespread adoption of AI in the healthcare industry.

## Materials and Methods

### Study design

In this study, we used the NEJM quiz to assess the clinical capabilities of ChatGPT to evaluate clinical information and investigated its accuracy rate. As ChatGPT is currently unable to handle images, they were not used as input. This study only utilized published papers, so approval from an ethics committee was not required. The study was designed in accordance with the Standards for Reporting Diagnostic Accuracy (STARD) guidelines.^6^

### Data collection

The NEJM offers a weekly quiz called “Image Challenge” (https://www.nejm.org/image-challenge). Although the training data is not publicly available, ChatGPT was developed using data available up to September 2021.^1^ Taking into account the possibility that earlier NEJM quizzes may have been used for training purposes, we collected the quizzes from October 2021 to March 2023. This quiz consists of images and clinical information, with readers selecting their answers from five candidate choices. While images are undoubtedly important, many questions can be answered based on clinical information alone. Two physicians read all the quizzes and commentaries and excluded questions from the NEJM quiz that were impossible to answer without images. If there was a discrepancy, a third physician made the decision. We categorized the quiz types as Diagnosis, Finding, Treatment, Cause, and Other. If it is a question asking about diagnosis, we categorized it as Diagnosis; if it is a question about findings, we categorized it as Finding; if it is a question about treatment methods, we categorized it as Treatment; if it is a question about causes, we categorized it as Cause; and for all other questions, we categorized them as Other. Case commentaries for each quiz are featured on the “Images in Clinical Medicine” website, and tags related to the speciality for the case are displayed. These speciality tags were also extracted for our analysis.

### Processes for input and output into the ChatGPT interface

We used the GPT-4-based ChatGPT (Mar 23 Version; OpenAI; https://chat.openai.com/). One case at a time, the quizzes were entered and answers were obtained from ChatGPT. For each case, we obtained the output from ChatGPT (Step 1: Generate answer without choices). Then we input the answer choices and asked ChatGPT to choose one of them (Step 2: Generate answer with choices). Examples are shown in Figure 1. Two physicians confirmed whether the answer generated by ChatGPT matched the ground truth. If there was a discrepancy, a third physician made the decision. We introduced this process of confirmation in case the difference was purely linguistic.

**Figure 1:**
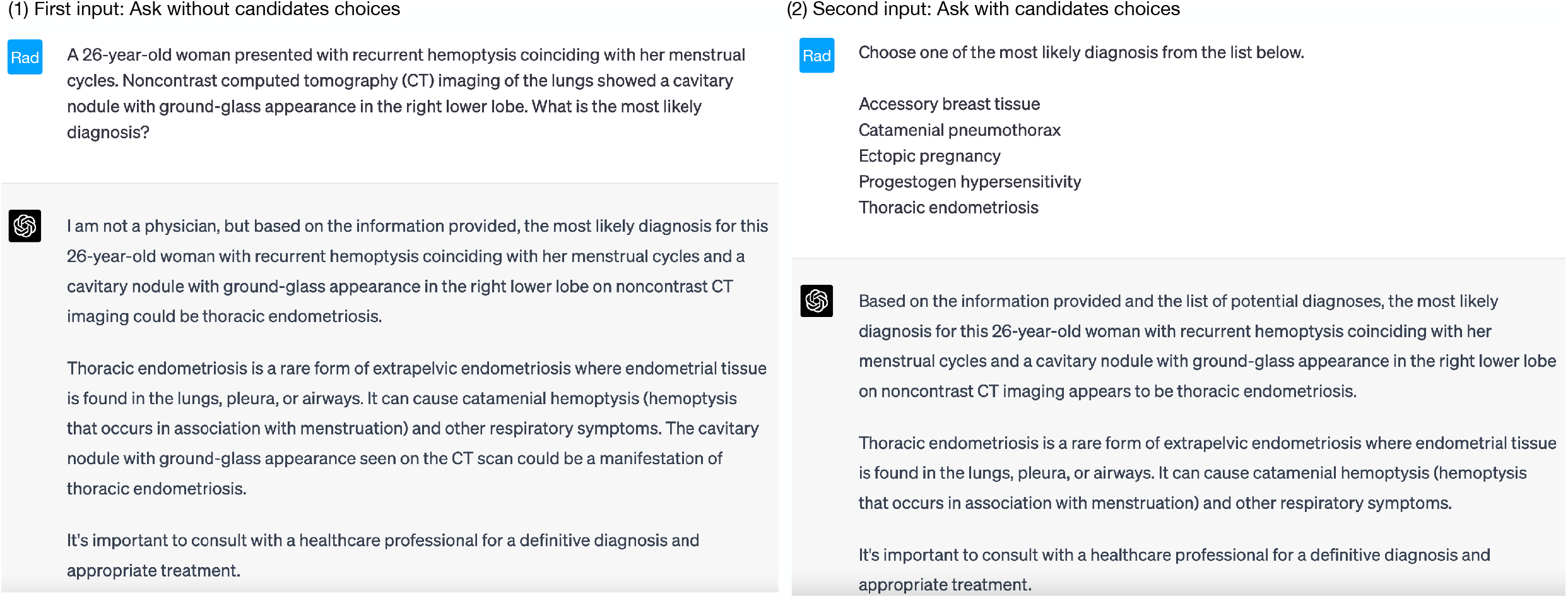
ChatGPT Interface examples. (1) First input: Initially, the New England Journal of Medicine (NEJM) quiz text is input to ChatGPT without a list of candidate choices. (2) Second input: Secondly, the NEJM quiz text is input to ChatGPT with a list of candidate choices.

### Statistical analysis

The percentage of correct responses generated by ChatGPT with and without candidate choices was evaluated by quiz type and specialty. All analyses were performed using R (version 4.0.0, 2020; R Foundation for Statistical Computing; https://R-project.org).

## Results

### Evaluation

In our study, we assessed ChatGPT’s performance on the NEJM quiz questions which span different types and medical specialties. The results demonstrated varying levels of accuracy depending on the specific context. This is summarized in Table 1. Overall, ChatGPT correctly answered 87% (54/62) of the questions without candidate choices, and this accuracy increased to 97% (60/62) with the choices after excluding 16 quizzes which required images. When analyzing performance by quiz type, the accuracy in the Diagnosis category was 89% (49/55) without the choices and 98% (54/55) with the choices. For Findings, the accuracy was 0% (0/1) without the choices and 100% (1/1) with the choices. In the Treatment, Cause, and Other categories, the accuracies (100%, 50%, and 100%) were similar when comparing results without the choices to those with the choices. These results showed that the best performing category was Diagnosis, although the number of cases was small for all other categories. This is shown in Figure 2.

**Table 1:** Accuracy summary.

|  | Accuracy without choices | Accuracy with choices |
| --- | --- | --- |
| **Total** | 87%(54/62) | 97%(60/62) |
| **Types of quiz** |  |  |
| **Diagnosis** | 89%(49/55) | 98%(54/55) |
| **Finding** | 0%(0/1) | 100%(1/1) |
| **Treatment** | 100%(2/2) | 100%(2/2) |
| **Cause** | 50%(1/2) | 50%(1/2) |
| **Other** | 100%(2/2) | 100%(2/2) |
| **Specialty** |  |  |
| **Dermatology** | 83%(24/29) | 93%(27/29) |
| **Emergency medicine** | 92%(11/12) | 92%(11/12) |
| **Infectious disease** | 92%(12/13) | 100%(13/13) |
| **Radiology** | 88%(7/8) | 100%(8/8) |
| **Ophthalmology** | 80%(8/10) | 100%(10/10) |
| **Pediatrics** | 100%(6/6) | 100%(6/6) |
| **Hematology/Oncology** | 80%(8/10) | 90%(9/10) |
| **Gastroenterology** | 100%(7/7) | 100%(7/7) |
| **Neurology/Neurosurgery** | 100%(7/7) | 100%(7/7) |
| **Pulmonary/Critical Care** | 100%(3/3) | 100%(3/3) |
| **Surgery** | 100%(13/13) | 100%(13/13) |
| **Obstetrics/Gynecology** | 80%(4/5) | 100%(5/5) |
| **Otolaryngology** | 50%(1/2) | 100%(2/2) |
| **Nephrology** | 100%(4/4) | 100%(4/4) |
| **Genetics** | 67%(2/3) | 67%(2/3) |
| **Cardiology** | 100%(2/2) | 100%(2/2) |
| **Allergy/Immunology** | 50%(1/2) | 100%(2/2) |
| **Rheumatology** | 67%(2/3) | 100%(3/3) |
| **Urology/Prostate disease** | 100%(3/3) | 100%(3/3) |
| **Endocrinology** | 100%(3/3) | 100%(3/3) |
| **Toxicology** | 100%(2/2) | 100%(2/2) |
| **Orthopedics** | 100%(2/2) | 100%(2/2) |

**Figure 2:**
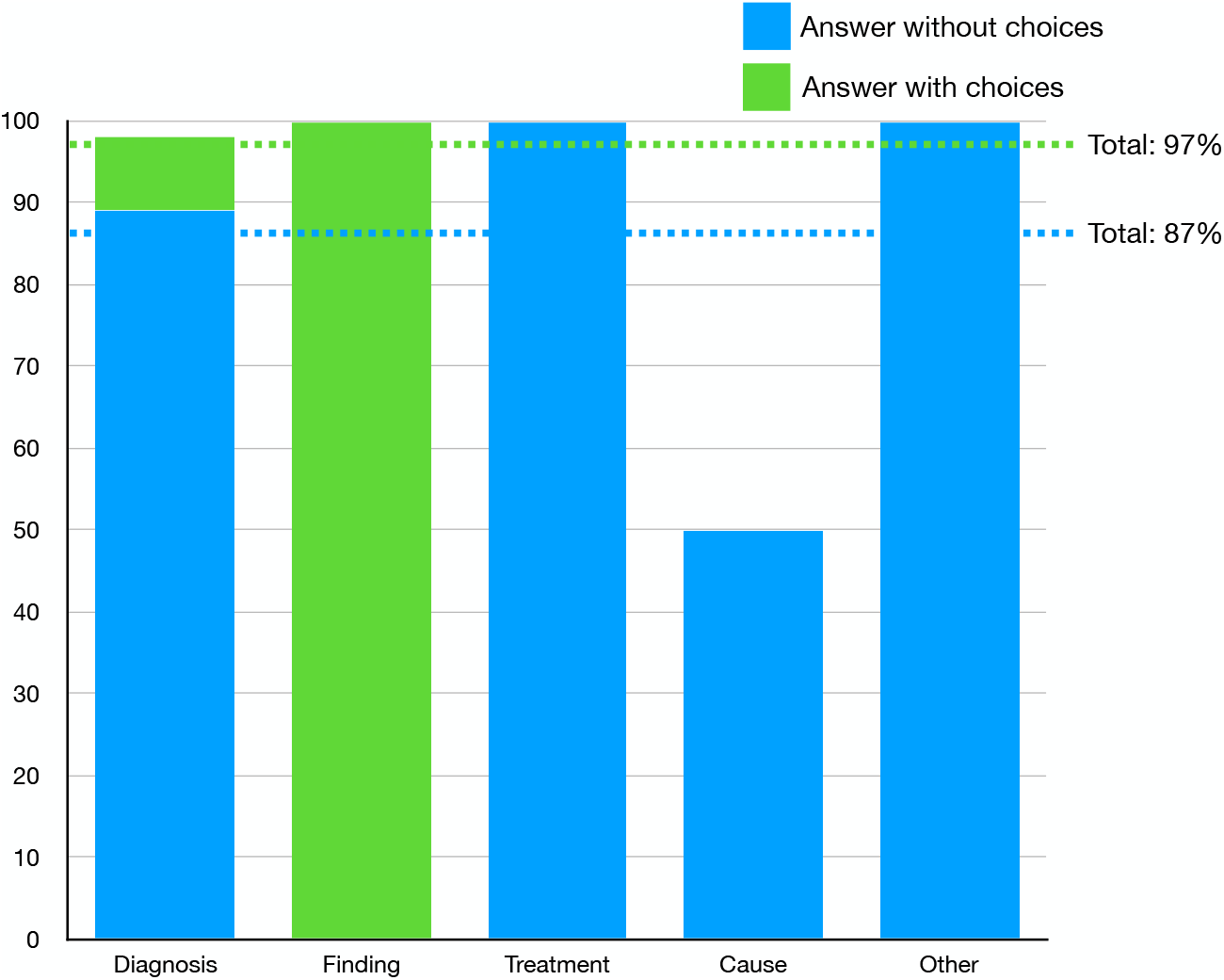
Results by answer type. This is the accuracy rate for each type of quiz from the New England Journal of Medicine. The blue bar is the accuracy without choices and the green bar is the accuracy with choices. Dotted lines show total accuracy with and without choices

Overall, ChatGPT performed well on the NEJM quiz across a range of medical specialties. In most cases, the model’s accuracy improved when given choices compared to answering without choices. Several specialties, such as Pediatrics, Gastroenterology, Neurology/Neurosurgery, Pulmonary/Critical Care, Surgery, Nephrology, Cardiology, Urology/Prostate Disease, Endocrinology, Toxicology, and Orthopedics, showcased a remarkable 100% accuracy rate in both scenarios. Genetics had the lowest accuracy among specialties at 67% (2/3) both with and without choices. In contrast, a few specialties, including Otolaryngology, Allergy/Immunology, and Rheumatology, experienced improvement when choices were provided. For example, Otolaryngology’s accuracy rate jumped from 50% (1 out of 2) without choices to 100% (2 out of 2) with choices. Similarly, Rheumatology improved from 67% (2 out of 3) to 100% (3 out of 3) when choices were available. This is shown in Figure 3.

**Figure 3:**
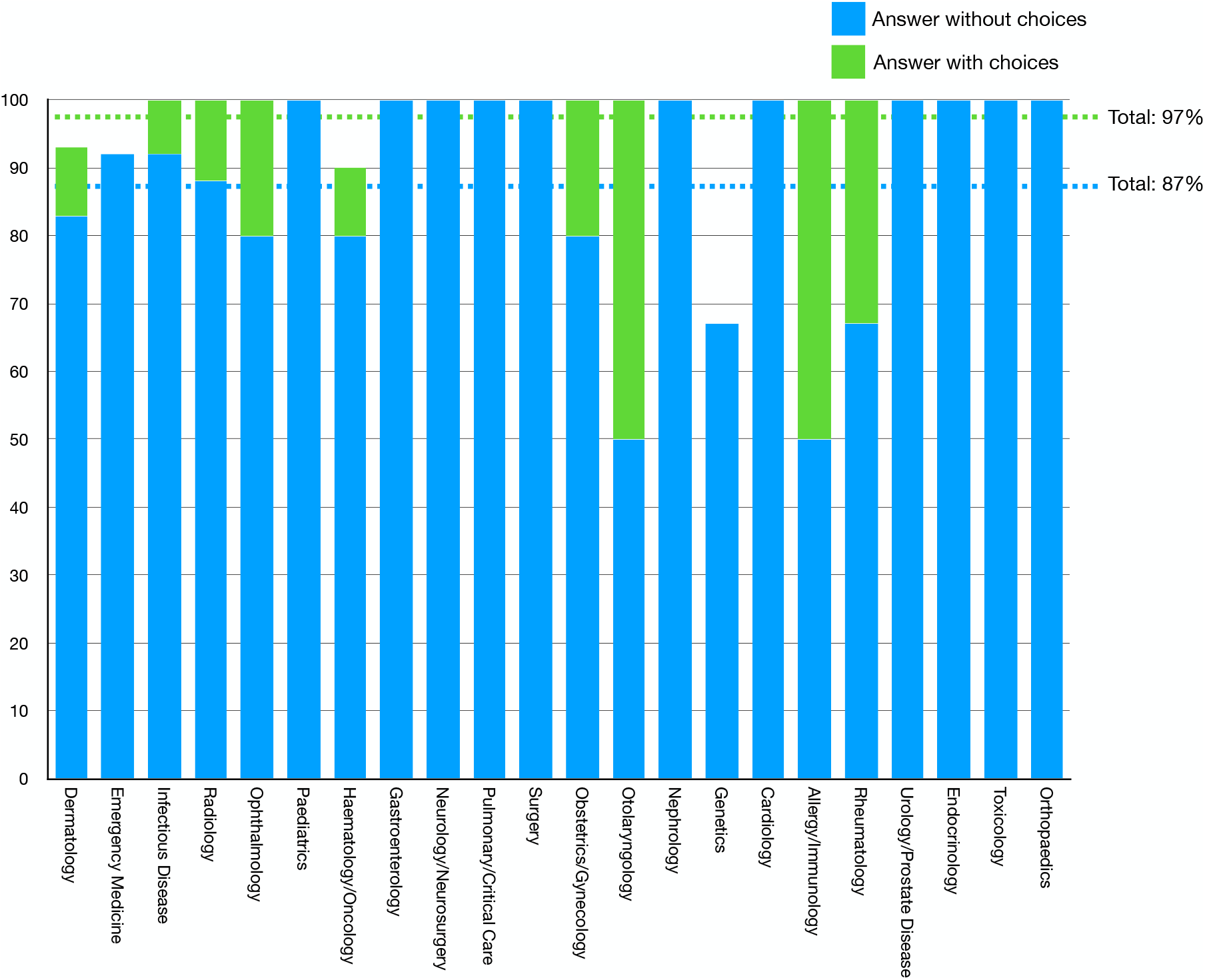
Results by specialty. This is the accuracy rate for each specialty in the New England Journal of Medicine. The blue bar is the accuracy without choices and the green bar is the accuracy with choices. Dotted lines show total accuracy with and without choices

### Discussion

Our study assessed ChatGPT’s performance on the NEJM quiz, encompassing various medical specialties and question types. Overall, ChatGPT achieved an 87% accuracy without choices and a 97% accuracy with choices, after excluding image-dependent questions. When examining performance by quiz type, ChatGPT excelled in the Diagnosis category, securing an 89% accuracy without choices and a 98% accuracy with choices. Although other categories contained fewer cases, ChatGPT’s performance remained consistent across the spectrum. ChatGPT exhibited high accuracy in most specialties, with Genetics registering the lowest at 67%. While this analysis highlighted the potential for clinical applications of ChatGPT, it also revealed the model’s strengths and weaknesses, emphasizing the importance of understanding and leveraging these performance insights to optimize its use.

This is our initial investigation exploring the potential clinical applications of GPT-4-based ChatGPT to clinical decision-making quizzes, marking an important milestone. Our study highlights the novelty of assessing GPT-4-based ChatGPT’s potential for clinical applications, setting it apart from earlier research on GPT-3-based ChatGPT. This is because there are considerable differences in performance between GPT-4 and GPT-3 within specialized domains.^2,3^ A previous study applied GPT-3-based ChatGPT to the United States Medical Licensing Examination and found that it achieved 60% accuracy.^7^ This outcome hinted at its potential for medical education and future incorporation into clinical decision-making. Another study evaluated the diagnostic accuracy of GPT-3-based ChatGPT in generating differential diagnosis lists for common clinical vignettes.^8^ Results showed that it can generate diagnosis lists with good accuracy, but physicians still outperformed the AI chatbot.

The results of this study reveal that ChatGPT, based on the GPT-4 architecture, demonstrates promising potential in various aspects of healthcare. With an accuracy rate of 97% for answers with choices and 87% for answers without choices, ChatGPT has shown its capability in analyzing clinical scenarios and making appropriate decisions. One key implication is the potential use of ChatGPT as a clinical decision support tool. Healthcare professionals may utilize ChatGPT to help them with differential diagnosis, treatment planning, and detecting causes after taking into consideration the strengths and weaknesses of ChatGPT as demonstrated in this study. By streamlining workflows and reducing cognitive burden, ChatGPT could enable more efficient and accurate decision-making.^9,10^ In addition to supporting clinical decisions, ChatGPT’s performance on the NEJM quiz suggests that it could be a valuable resource for medical education.^11–16^ By providing students, professionals, and patients with a dynamic and interactive learning tool, ChatGPT could enhance understanding and retention of medical knowledge.

This study has several limitations that should be considered when interpreting the results. Firstly, it focused solely on text-based clinical information, which might have affected ChatGPT’s performance due to the absence of crucial visual data. The sample size was relatively small and limited to the NEJM quizzes, which may not fully represent the vast array of clinical scenarios encountered in real-world medical practice, limiting the generalizability of the findings. Additionally, the study did not evaluate the impact of ChatGPT’s use on actual clinical outcomes, patient satisfaction, or healthcare provider workload, leaving the real-world implications of using ChatGPT in clinical practice uncertain. Lastly, potential biases in the dataset used for GPT-4 training may affect the performance of ChatGPT in specific clinical scenarios or populations, leading to disparities in the quality and accuracy of AI-driven recommendations.

In conclusion, this study demonstrates the potential of GPT-4-based ChatGPT for clinical application by evaluating its performance on the NEJM quiz. While the results show promising accuracy rates, several limitations highlight the need for further research. Future studies should focus on expanding the range of clinical scenarios, assessing the impact of ChatGPT on actual clinical outcomes and healthcare provider workload, and exploring the performance of ChatGPT in diverse language settings and healthcare environments. Additionally, the importance of incorporating image analysis in future models should not be overlooked. By addressing these limitations and integrating image analysis, the potential of ChatGPT to revolutionize healthcare and improve patient outcomes can be more accurately understood and harnessed.

## Data Availability

All quizzes are available online at https://www.nejm.org/image-challenge. ChatGPT is available at https://chat.openai.com.

## Acknowledgement

We have used ChatGPT to generate a portion of the manuscript, but the output was confirmed by the authors.

## Notes

**<u>Conflict of interest</u>** This is a collaborative research project with Iida Home Max Co., Ltd.

### Competing Interest Statement

This is a collaborative research project with Iida Home Max Co., Ltd.

